# Modeling and Dynamics in Epidemiology, COVID19 with Lockdown and Isolation Effect : Application to Moroccan Case

**DOI:** 10.1101/2020.04.29.20084871

**Authors:** Radouane Yafia

**Affiliations:** Ibn Tofail University

**Keywords:** SIR model, Covid19, local stability, lockdown, isolation, basic reproduction number *R*_0_.

## Abstract

In this paper, we present a SIR mathematical model in order to study the dynamics and propagation of Covid19 and the effect of lockdown of susceptible population and isolation of infectious population. The basic reproduction number *R*_0_ depends on the lockdown and isolation rates. We prove that, *R*_0_ becomes smaller than one if the lockdown and isolation rates are higher and in this case, we have the extinction of the infectious population. We apply our results to the case of Morocco country. In the end we carried out some numerical simulations illustrating our results.

## Remark 1

All the figures in this paper are obtained from simplified system of differential equations and only give predictions up to the peak of the disease, these approximations are only reliable if t < t_peak_. For t > t_peak_ one cannot reasonably predict the evolution of the epidemic, (where t is the time, t_pea_k indicates when the peak of the epidemic occurs). All our numerical results depend on rates of transmission, cure, lethality, lockdown and isolation, and incubation period, which cannot be accurately determined since they change from day to day.

## 1 Mathematical model

In this paper, a SIR model is proposed which is an extension of the classical Kermack and McKendrick model [4, 5, 6, 1], taking into account the effect of lockdown of susceptible and isolation of infectious populations. It is also assumed that 0 ≤ *α* < 1 and 0 ≤ *δ* < 1 are the lockdown rate of susceptible and isolation rate of infectious, respectively, so the fraction of susceptible that is contained (protected) is (1 − *α*)*S* and the fraction of non-isolated infectious population is (1 − *δ*)*I*. *Lambda* is the density of susceptible births. R population contains individuals removed from the transmission chain, the contained individuals given by *αS* proportion, the isolated individuals given by tôI proportion, the cured and the deceased. The mathematical model is given by:

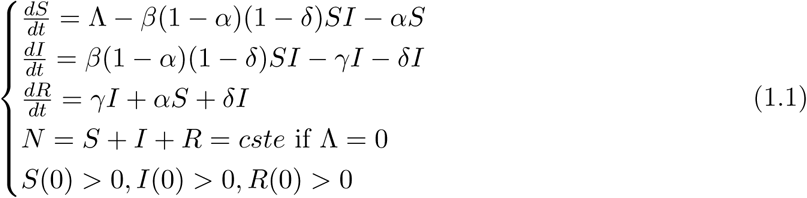

## 2 Covid19, March 2 – 8, Moroccan Case

In the following we consider the beginning of the disease (March 2 – 8) which corresponds to the case where the number of births is relatively negligible, Λ = 0, and there is no lokcdown, *α* = *δ* = 0. In this case it is assumed that *S*(*t*) = *S*_0_ and that the infected cases are only the imported cases, with zero cure and mortality, i.e. *γ* = 0. System (4.1) is then written as follows

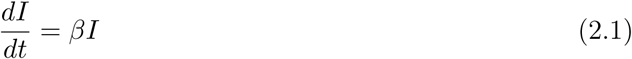

The solutions of (2.1) are of the form *I*(*t*) = *I*_0_*e^βt^*. After fitting the curve (Fig. 9 (a)), we obtain the increase rate is *β* = 0.17. At the beginning of the disease around March 2 the first case detected (imported from Italy) at March 8 the infection rate was of order of 0.17, the epidemic thus tends to grow exponentially, without mortality and without cure, in this case the doubling time is of order 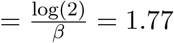, which is the necessary time for the number of infected population to double.

## 3 Covid19, March 8 – 19, Moroccan Case

We are now considering the second phase of the epidemic, and taking into account the period March 8 – 19. By linear regression (see Fig. 9 (c)), we find that the increase rate of infected population is equal to λ = 0.146, which corresponds to increase rate of *e^λt^*. During this period the doubling time is of order 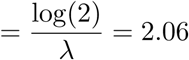. During this period, the epidemic grows than the first one. It’s assumed that the number of births is negligible, lambda=0, and that lokcdown is not yet applied, *alpha* = *delta* = 0.

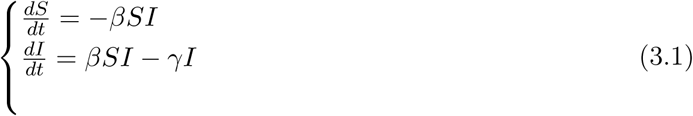

The equilibria of system (3.1) are (0,0) and (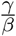, 0).

In this

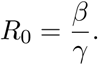

We see that if *R*_0_ > 1, we have 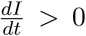, which implies that the number of infected population increases, and conversely if not, it decreases. Now let’s evaluate the final size of the susceptible population *S*(∞). By integrating the first and second equations between 0 and ∞, we have 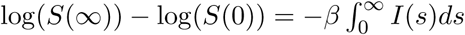 and 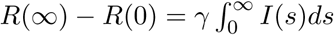.

As *R*(0) = 0, *S*(0) = 1 at the beginning of the epidemic and by a simple calculation we have

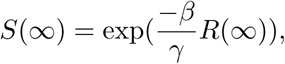

and

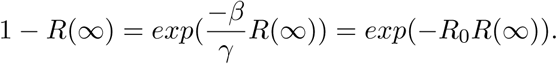

To get the equation in *S*(∞), let’s divide the second equation of (3.1) by the first one, we get

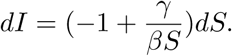

By a simple calculation, we have

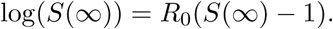

If *R*_0_ *>* 1, this equation has two solutions, one of which is between 0 and 1.

To estimate the transmission rate *β* along this period, we integrate the first equation between 8 and 19 days (see the table of statistics, Moroccan case), we obtain

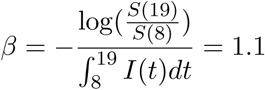

The estimation of 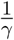 is given by the average time in the *I* compartment before isolation. It’s assumed it’s worth about 1 and

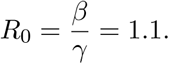

Since *R*_0_ *>* 1, but close to 1; this explains the low strength of infection at the beginning of the disease.

With a population of *N* = 340000 and from figure 1, we can see that from these approximations, the peak arrives 61 days after the beginning of the disease. Certainly, the number of infected population would be equal to approximately *I*(61) = 0.0229292 × 34000000 = 779280, the number of recovered population would be 0.1829 × 34000000 = 6218600 and the number of uninfected people would be 0.7929 × 34000000 = 26958600 with the assumption that no action of lockdown or isolation has been taken.

**Figure 1:**
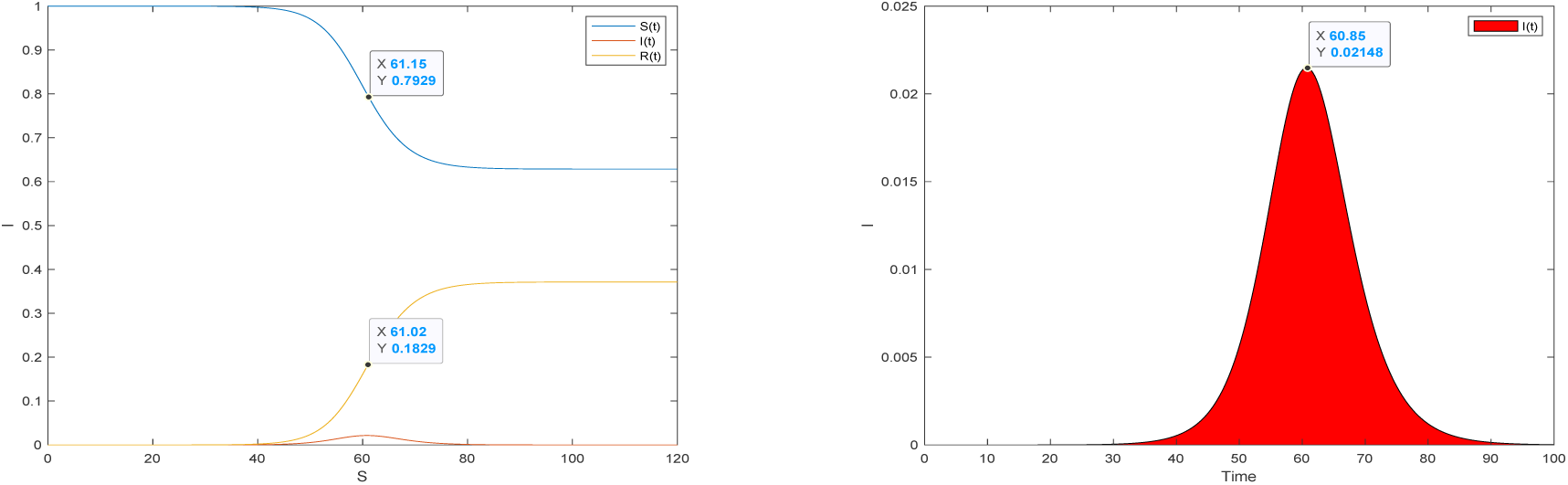
Temporal evolution of the solutions obtained from (3.1), with *β* = 1.25, *γ* = 1. These solutions are converging towards the equilibrium point (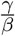, 0). These two figures give the daily number of infected population for *β* = 1.25 and *γ* = 1 and without lockdown.

**Figure 2:**
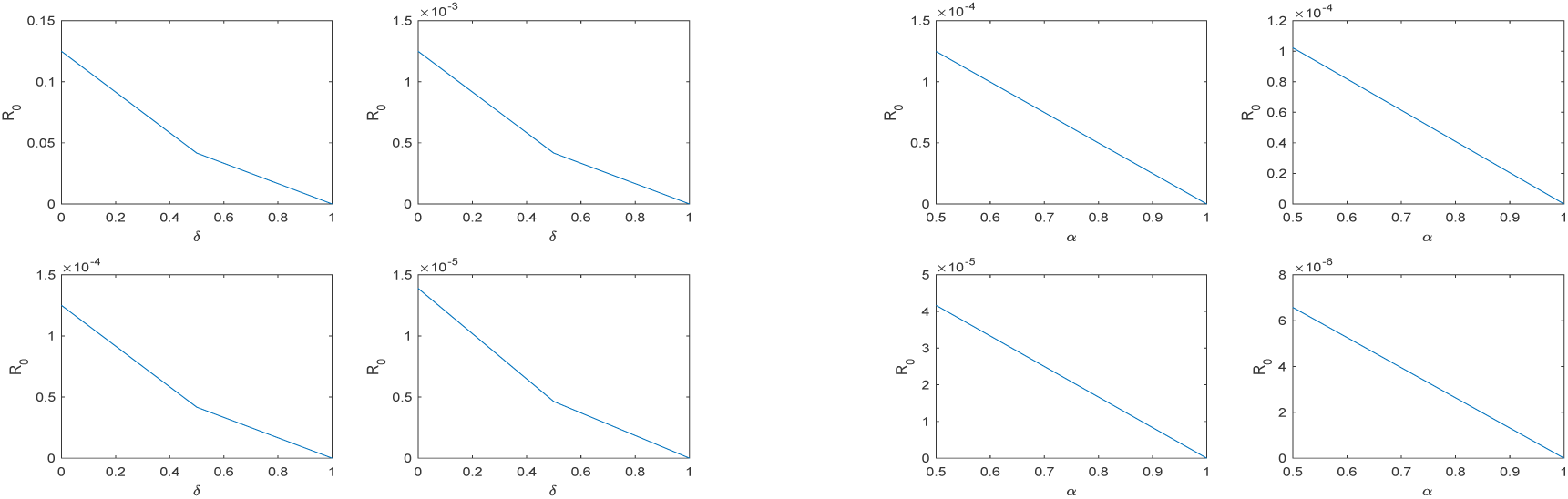
Variations of 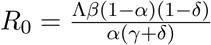 depending on *δ* with *α* = 0.0001, 0.1, 0.5, 0.9 (on the left), and depending on *α* with *δ* = 0.0001, 0.1, 0.5, 0.9 (on the right), for *γ* = 1, *β* = 1.25, Λ = 0.0001. We can see that, more *α* or *δ* increases more *R*_0_ decreases.

**Figure 3:**
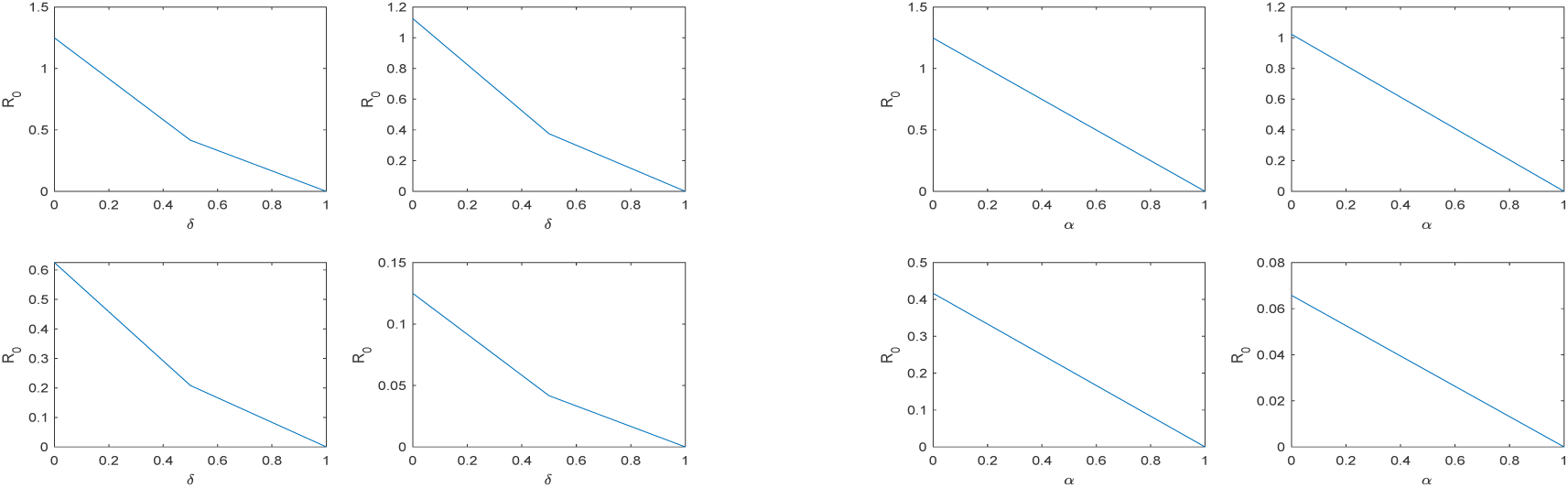
Variations of 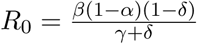 versus *δ* with the values of *α* = 0.0001, 0.1, 0.5, 0.9 (left), and as a function of *α* with the ‘values of *δ* = 0.0001, 0.1, 0.5, 0.9 (right) for *γ* = 1, *β* = 1.25. We can see that as *α* or *δ* increases, *R*_0_ decreases.

## 4 Disease progression with lockdown

### 4.1 Positivity, bounded solutions and equilibria

In this paragraph, we consider the model (1.1) with lockdown *α* > 0, and isolation *δ* > 0. As *R* population depends on *S* and *I* populations and that the latter are independent of *R*, we restrict our study to the *SI* system.

As the the disease take more times, we take into account the effect of new births. So the system becomes

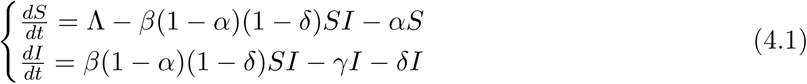

#### Proposition 2

*1-Any solution of system* (4.1) *starting from a positive initial condition remains positive for any t >0. 2-Any solution of system* (4.1) *is bounded*.

Equilibria of (4.1):

The equilibrium points are obtained by solving the algebraic system obtained by cancelling all derivatives of *S*(*t*) and *I*(*t*). Thus:

1- The Disease free equilibrium (DFE) is: 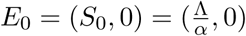.

2- The endemic equilibrium (EQ) is: 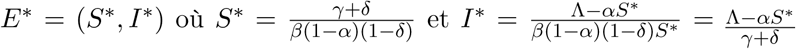.

The endemic equilibrium *E*\* is positive if 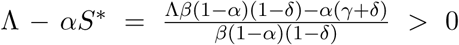, which imply 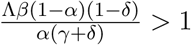.

Let

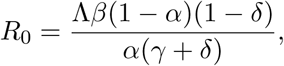

the basic reproduction number (rate). It can also be calculated by the “next generation matrix” method introduced by Diekmann and Heesterbeek (voir [2]).

### 4.2 Stability of equilibria

#### Proposition 3

*Suppose R_0_ <1, so:*

1- *The disease-free equilibrium is the only equilibrium of of system 4.1. 2- The equilibrium point E_0_ is locally asymptotically stable*.

##### Proof

Let *E* = (*S,I*) be an equilibrium point of system 4.1, by linearization we obtain the following jacobian matrix:

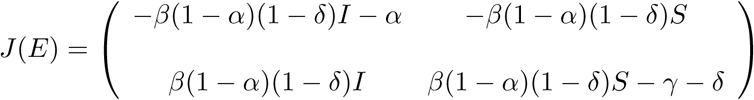

Thus, the Jacobian matrix associated with *E*_0_ is

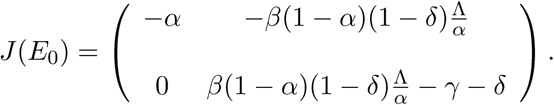

The characteristic equation is defined by:

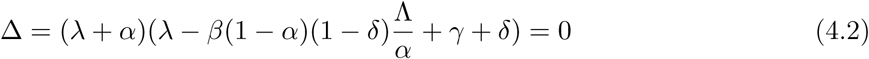

and the corresponding eigenvalues are *λ*_1_ = −*α* < 0, 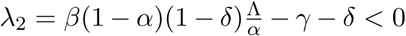. Hence, we deduce the result.

#### Proposition 4

*Suppose R*_0_ *>1, so:*

1- *The disease free equilibrium E*_0_ *of system 4.1 is unstable*.

2- *The endemic equilibrium point E*\* *of system 4.1 is locally asymptotically stable*.

##### Proof

1- Obvious..

2- The Jacobian matrix associated with *E*\* is given by

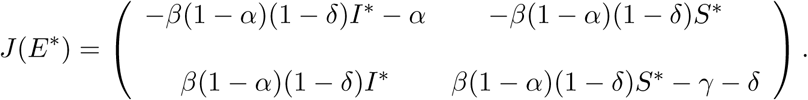

As 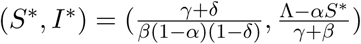 et the jacobienne becomes

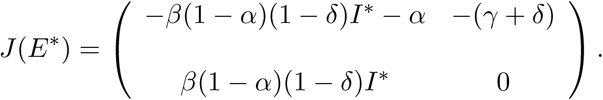

The characteristic equation is given by:

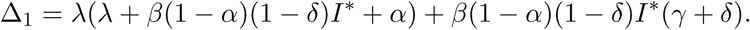

As

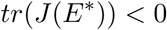

and

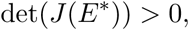

the endemic equilibrium *E*^*^ is locally asymptotically stable.

### Remark 5

*On the other hand, it can also be shown that the endemic equilibrium E*\* *is globally asymptotically stable using an adequate Lyapunov function*.

### Remark 6

*Turning off an epidemic such as Covid19, is like controlling the basic reproduction number R*_0_ (*i.e. making it strictly less than 1*)*. According to the formula of R*_0_, *if we increase the lockdown rate α of susceptible population and the isolation rate δ of infected population* (*reported or not*)*, we can make R*_0_ < 1, *the number of infected population thus decreasing towards the extinction point. On the other hand, if R*_0_ *remains greater than* 1, *the number of infected population increases day by day and the disease persists over time, since the endemic point, in this case, is globally asymptotically stable*.

### Remark 7

*If now we assume that the total population is constant during the outbreak* Λ = 0, *in this case system* (4.1) *is written as*

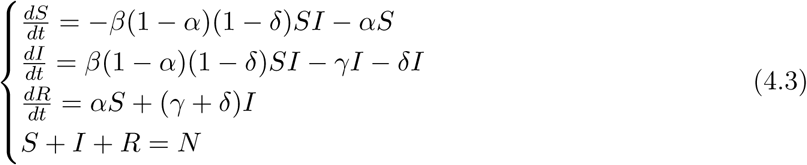

*and* 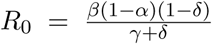. *With the previous values and for different values of α and δ, we give simulations that take into account the role of lockdown of susceptibles and the isolation of infectious population of system* (4.3)*. According to Fig 9* (*b*), *we observe that the increase rate of infected after lockdown is equal to* 0.0673, *which is smaller than the increase rate before, implying that the transmission rate becomes smaller, and we then take β* = 1.1 *instead of* 1.25.

## 5 Conclusions

In conclusion, in this article it was possible to determine the role of lockdown of susceptible population and isolation of infectious population, depending on the basic reproductive number, which is a key element in the spread of epidemics in general, and of Covid19 in particular. To deal with this problem we have subdivided the time period from 2 March to 5 April 2020 into three periods:

1- Beginning of the outbreak (March 2–8): Appearance of a few imported cases.

2- Intermediate period (March 8–19): local cases appear.

3- Period after containment (March 20…): spread of epidemic within population.

We showed that, in the first period, the transmission rate was low, which explains the moderate growth in the cumulative number of infected population. In the second period, the transmission rate of the epidemic becomes higher, and the cumulative number of infected population increases. In the last phase and taking into account lockdown and isolation, the transmission rate becomes smaller compared to the second phase. Since the basic reproductive number depends on the lockdown and isolation rates, we have shown that if these rates are increased, the number becomes smaller than 1, and larger than 1 otherwise. In conclusion, controlling an outbreak means controlling the lockdown rate of the susceptible and isolation rate of infected, which is confirmed by figures 4, 5 and 6.

**Figure 4:**
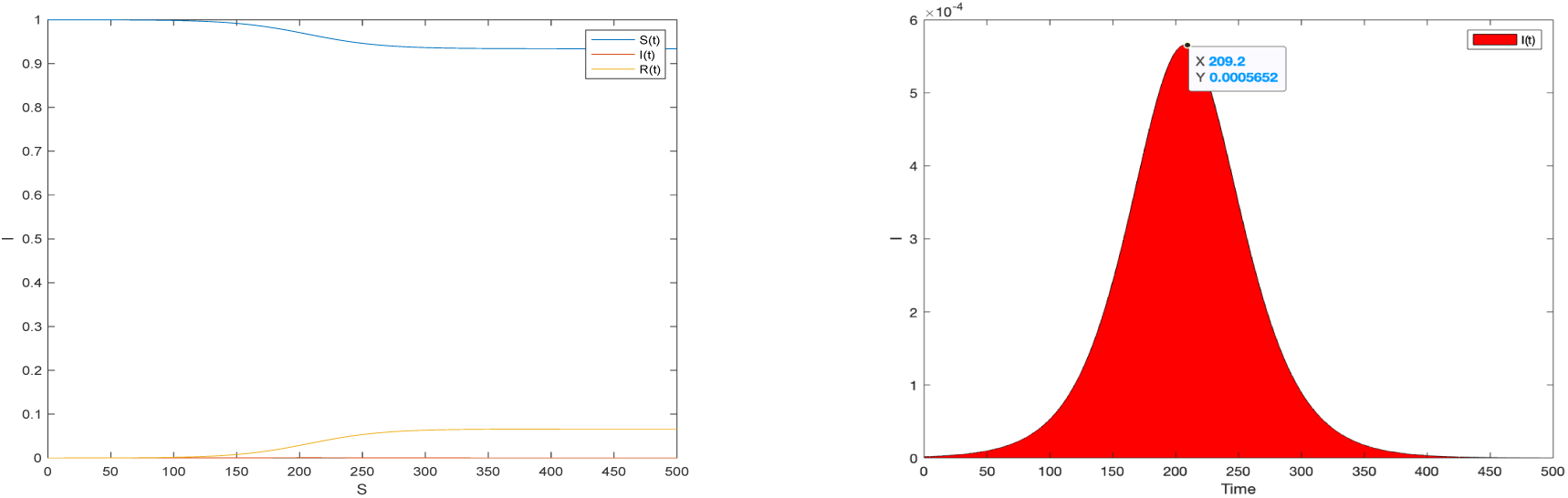
Time evolution of solutions of system (4.3) *R*_0_ = 1.243 for *α* = 0.05, *δ* = 0.01, *γ* = 1, *β* = 1.1, the peak is reached at 19216, a much lower value with a multiplicative factor of about 50 than in the case without lockdown, after 209 days. In the figure on the left, the inference curve is so low that it seems to be confused with the abscissa axis, as can be seen in the figure on the right.

**Figure 5:**
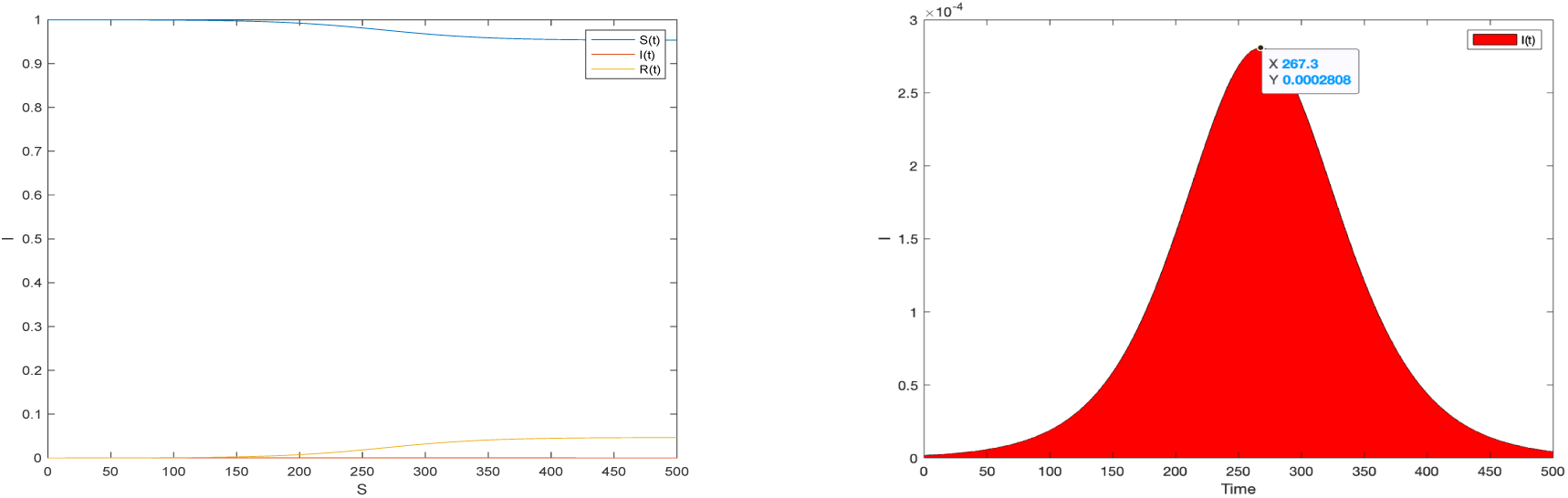
Temporal evolution of solutions of system (4.3) *R*_0_ = 1.004 for *α* = 0.05, *δ* = 0.02, *γ* = 1, *β* = 1.1, the peak is reached at 9547, a much lower value with a multiplicative factor of order of about 50 than in the case without lockdown, after 267 days. In the figure on the left, the inference curve is so low that it seems to be confused with the abscissa axis, as can be seen in the figure on the right.

**Figure 6:**
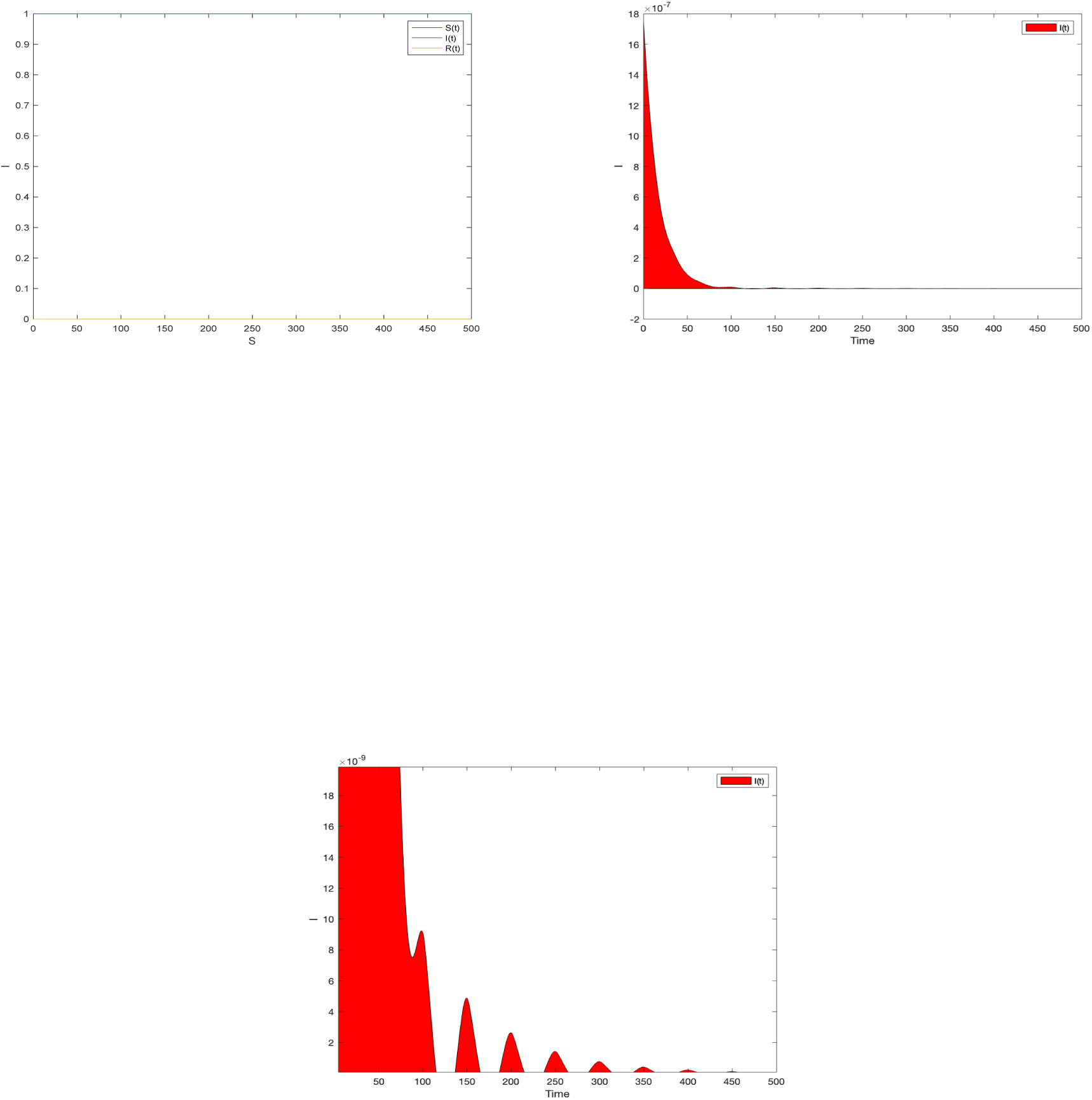
Temporal evolution of solutions of system (4.3) *R*_0_ = 0.885 for *α* = 0.05, *δ* = 0.05, *γ* = 1, *β* = 1.1, if the lockdown is very strict the number of infected people decreases towards 0, nevertheless there is an appearance of a succession of peaks lower and lower, disappearing with time.

**Figure 7:**
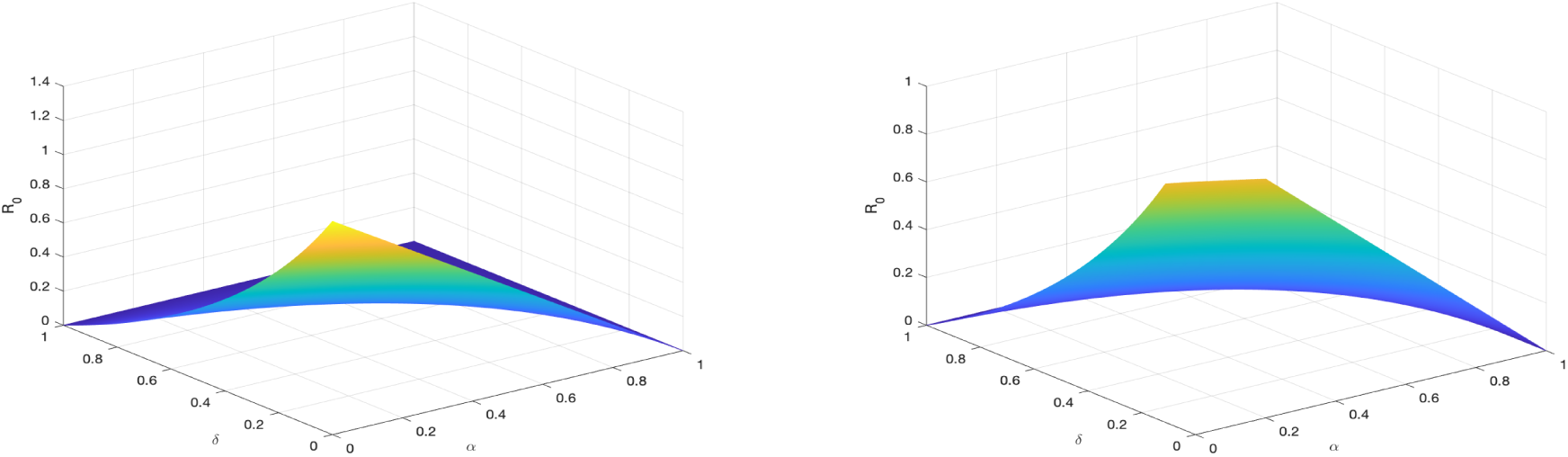
For *γ* =1; *β* = 1.1, variations of 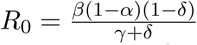 depending on *α* et *δ*: from a certain value of these parameters *R*_0_ becomes < 1.

**Figure 8.**
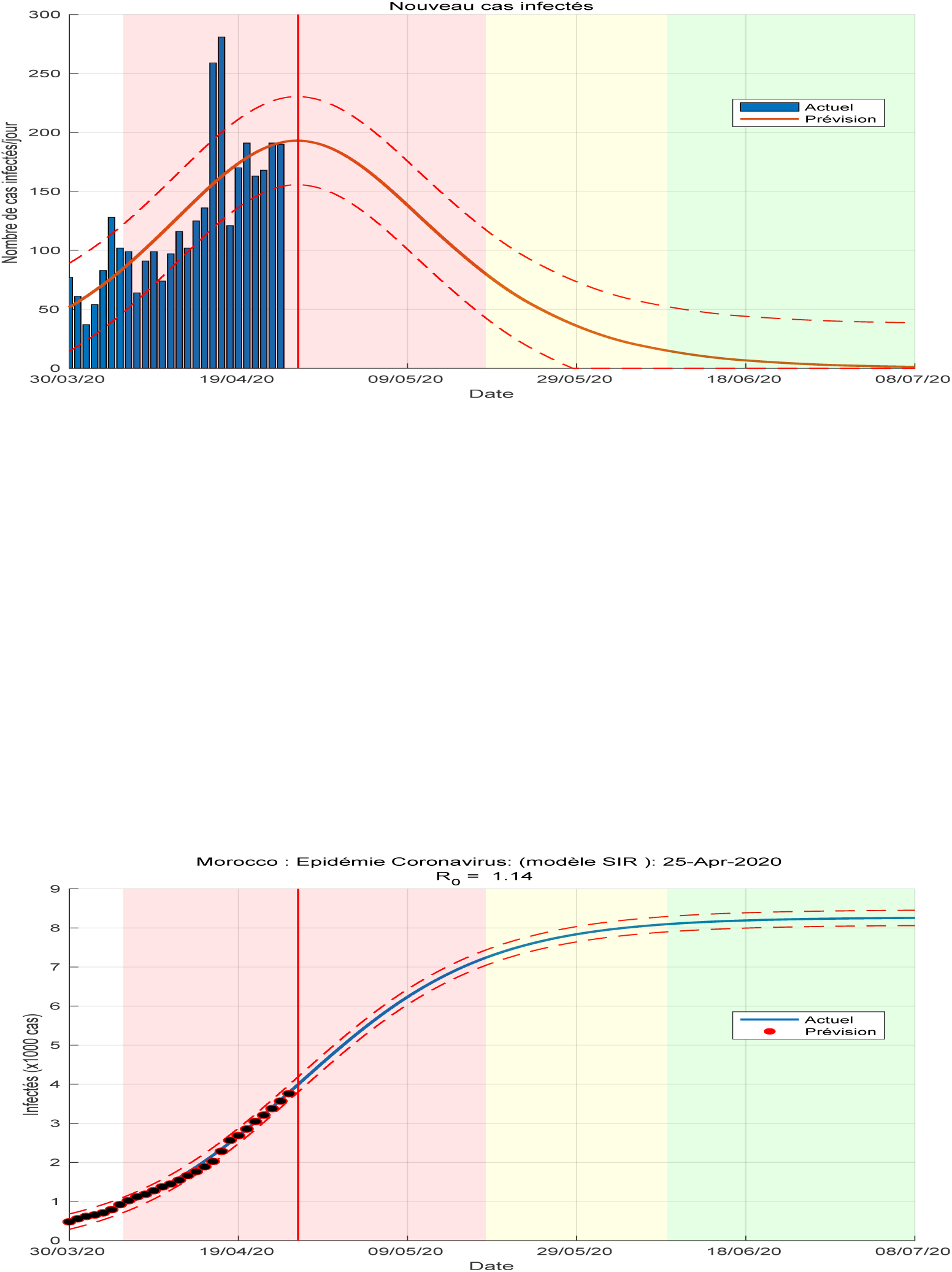

**Figure 9:**
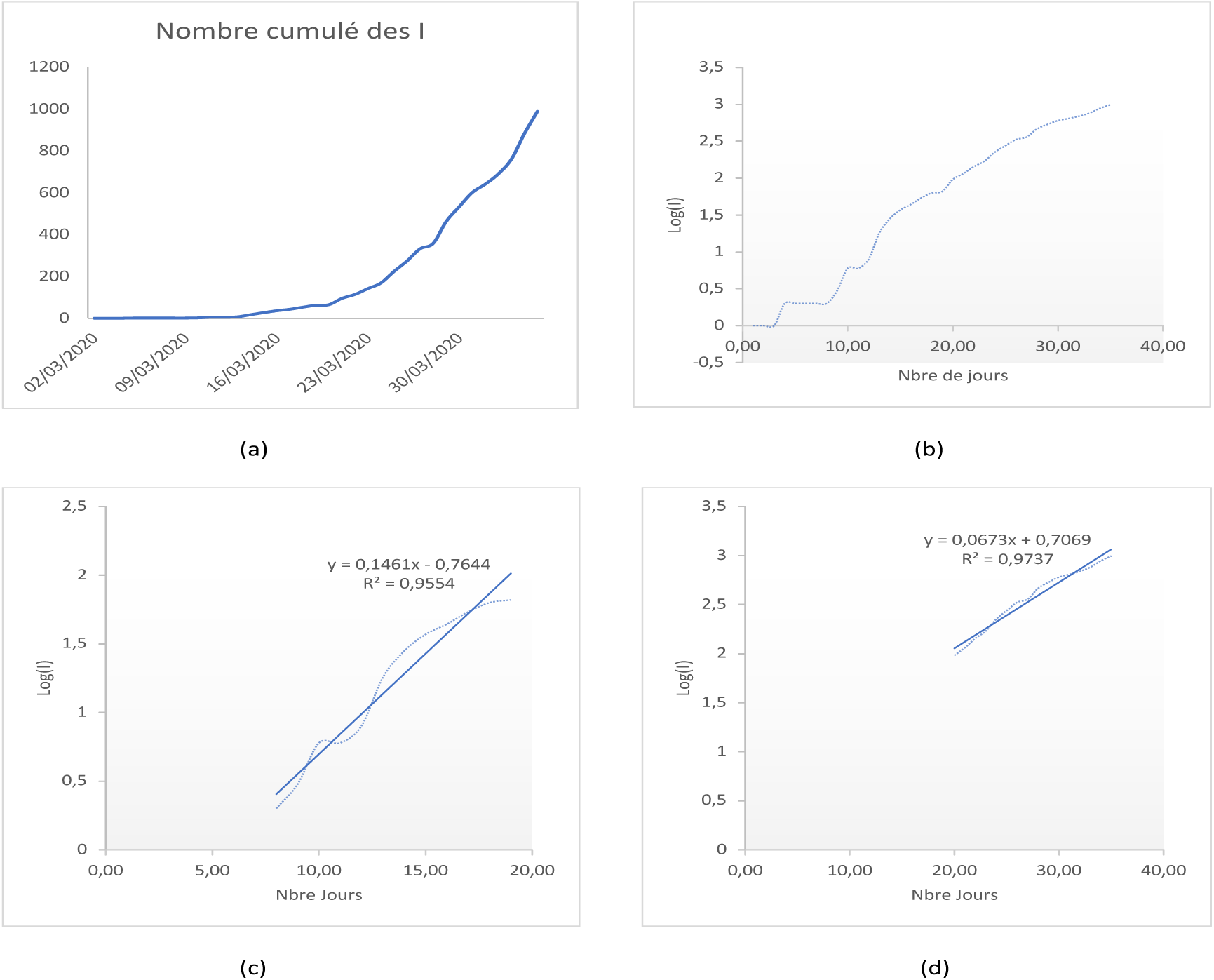
(a): Cumulative number of infected cases. (b): Logarithm of cumulative number of infected cases, (c): Linear regression calculated over the period March 8 to March 19 without lockdown and isolation, (d): Linear regression calculated over the period from March 20 to April 5 after lockdown and isolation. These figures are based on statistics reported by the Ministry of Health, Morocco.

## 6 Forecast: Morocco

We end our article with a forecast for Moroccan country. We use the SIR model, by plotting the cumulative and daily number of infected population from March 12 2020 [7]

## Data Availability

Ministry of Health Mrorcco

## Acknowledgements

I would like to thank Prof. Aziz Alaoui from the University of Le Havre for his proofreading and his remarks which allowed me to improve the quality and the writing of this paper. I also thank

